# Clinical course impacts early kinetics and long-term magnitude and amplitude of SARS-CoV-2 neutralizing antibodies beyond one year after infection

**DOI:** 10.1101/2021.08.12.21261921

**Authors:** Edwards Pradenas, Benjamin Trinité, Víctor Urrea, Silvia Marfil, Ferran Tarrés-Freixas, Raquel Ortiz, Carla Rovirosa, Jordi Rodon, Júlia Vergara-Alert, Joaquim Segalés, Victor Guallar, Alfonso Valencia, Nuria Izquierdo-Useros, Marc Noguera-Julian, Jorge Carrillo, Roger Paredes, Lourdes Mateu, Anna Chamorro, Ruth Toledo, Marta Massanella, Bonaventura Clotet, Julià Blanco

**Affiliations:** IrsiCaixa AIDS Research Institute, Germans Trias i Pujol Research Institute (IGTP), Can Ruti Campus, UAB, 08916, Badalona, Catalonia, Spain; IRTA Centre de Recerca en Sanitat Animal (CReSA, IRTA-UAB), Campus de la UAB, 08193, Bellaterra, Catalonia, Spain; UAB, Centre de Recerca en Sanitat Animal (IRTA-UAB), Campus de la UAB, 08193, Bellaterra, Catalonia, Spain; Departament de Sanitat i Anatomia Animals, Facultat de Veterinària, UAB, 08193, Bellaterra, Catalonia, Spain; Barcelona Supercomputing Center, 08034, Barcelona, Catalonia, Spain; Catalan Institution for Research and Advanced Studies (ICREA), 08010, Barcelona, Catalonia, Spain; Infectious Diseases Department, Fight against AIDS Foundation (FLS), Germans Trias i Pujol Hospital, 08916, Badalona, Catalonia, Spain; University of Vic–Central University of Catalonia (UVic-UCC), 08500, Vic, Catalonia, Spain

**Author notes:** **Corresponding author:** Julià Blanco, PhD, Senior Researcher, Institut de Recerca de la Sida. IrsiCaixa, IGTP, Hospital Germans Trias i Pujol, Ctra. de Canyet s/n. 2a Planta Maternal. 08916 Badalona. Barcelona. Equal contribution (Edwards Pradenas, Benjamin Trinité). **Funding** Grifols, Departament de Salut of the Generalitat de Catalunya, the Spanish Health Institute Carlos III, CERCA Programme/Generalitat de Catalunya, ANID, and the crowdfunding initiatives #joemcorono, BonPreu/Esclat and Correos.

**Keywords:** SARS-CoV-2, humoral response, B-cell memory, pseudovirus, neutralizing antibodies, variants of concern

## Abstract

**Background:** Understanding the determinants of long-term immune responses to SARS-CoV-2 and the concurrent impact of vaccination and emerging variants of concern will guide optimal strategies to achieve global protection against the COVID-19 pandemic.

**Methods:** A prospective cohort of 332 COVID-19 patients was followed beyond one year. Plasma neutralizing activity was evaluated using HIV-based reporter pseudoviruses expressing different SARS-CoV-2 spikes and was longitudinally analyzed using mixed-effects models.

**Findings:** Long-term neutralizing activity was stable beyond one year after infection in mild/asymptomatic and hospitalized participants. However, longitudinal models suggest that hospitalized individuals generate both short- and long-lived memory B cells, while outpatient responses were dominated by long-lived B cells. In both groups, vaccination boosted responses to natural infection, although viral variants, mainly B.1.351, reduced the efficacy of neutralization. Importantly, despite showing higher neutralization titers, hospitalized patients showed lower cross-neutralization of B.1.351 variant compared to outpatients. Multivariate analysis identified severity of primary infection as the factor that independently determines both the magnitude and the inferior cross-neutralization activity of long-term neutralizing responses.

**Conclusions:** Neutralizing response induced by SARS-CoV-2 is heterogeneous in magnitude but stable beyond one year after infection. Vaccination boosts these long-lasting natural neutralizing responses, counteracting the significant resistance to neutralization of new viral variants. Severity of primary infection determines higher magnitude but poorer quality of long-term neutralizing responses.

## Introduction

Immune responses to SARS-CoV-2 infection involve an undefined balance of innate and adaptive pathways^1^ resulting in the development of a seemingly long-lasting immunological memory.^2,3^ Although there is a general consensus on the key role of both T and B cells on protection against SARS-CoV-2 infection and development of COVID-19, the specific contribution of each arm of the immune system is still unclear.^1^ Neutralizing antibodies mediate their protective effect by binding to the spike (S) glycoprotein of SARS-CoV-2 and blocking viral entry into target cells; however, additional effector functions promoting viral clearance or natural killer (NK)-mediated infected-cell killing seems to be also relevant in SARS-CoV-2 and other viral infections.^4^ Nevertheless, abundant experimental and epidemiological work on SARS-CoV-2 indicate that neutralizing antibodies can serve as surrogate marker of protection,^5–7^ as for other viral infections.^8,9^

Given the relevance of antibodies, the early (1-3 months) and mid-term (3-12 months) humoral response after SARS-CoV-2 infection has been thoroughly described.^10–14^ Current data outline a heterogeneous scenario, in which infected individuals generate a wide range of neutralizing antibodies (from no seroconversion to rapid development of high titers) with no definitive association to age, gender or disease severity.^15–17^ Various authors have also suggested complex kinetics of neutralizing activity decay.^18–20^ This is particularly relevant in the current context of viral evolution, as several variants of concern (VOCs) have shown total or partial resistance to neutralizing antibodies and partial resistance to polyclonal humoral responses elicited by infection or vaccination.^21^

To understand the dynamics of natural responses to infection, we focused on the longitudinal analysis of the neutralizing humoral response in a large cohort of mild/asymptomatic and hospitalized individuals infected by SARS-CoV-2. Our analysis includes one of the longest follow-up periods (up to 15 months) and shows that the long-term magnitude of neutralization is remarkably stable, being boosted by vaccines and potentially threatened by VOCs. Clinical severity of primary infection was identified as the main factor determining the kinetics, the magnitude and the quality of neutralizing antibodies.

## Results

### Cohort description

Our cohort included 332 participants with confirmed SARS-CoV-2 infection, recruited between March 2020 and March 2021 in Catalonia (North-Eastern Spain). Participants were grouped according to the epidemiological waves of SARS-CoV-2 pandemic in Spain, defined by an early outbreak caused by the original Wuhan-Hu-1 (WH1) and the B.1 variant (D614G) (from March to June 2020), a second wave dominated by the 20E (EU1) variant (from July to December 2020), and a third wave associated with the emergence of the B.1.1.7 alpha variant covering the January to June 2021 period, until the recent introduction of the B.1.617.2 delta variant in June 2021 (Supplementary Figure 1). A total of 212 participants were recruited during the first wave period, 128 with mild or absent symptomatology (WHO progression scale^22^ levels 1-3; outpatients) and 84 having required hospitalization (WHO progression scale^22^ levels 4-10) with a wide range of severity from non-severe pneumonia to intensive care unit admission/death (Table 1). Comparable proportions of disease severity were observed in patients recruited in the second (n=79) and third (n=41) COVID-19 waves. In all cases, the hospitalization groups showed older age and lower female frequency when compared with outpatients (mild or asymptomatic, Table 1).

**Table 1.** Characteristics of individuals included in analysis

|  | March 20 – June 20 |  | July 20 – December 20 |  | January 21 – March 21 |  |  |
| --- | --- | --- | --- | --- | --- | --- | --- |
|  | Outpa-<br>tients<br>(n=128) | Hospita-<br>lized<br>(n=84) | Outpa-<br>tients<br>(n=43) | Hospita-<br>lized<br>(n=36) | Outpa-<br>tients<br>(n=19) | Hospita-<br>lized<br>(n=22) | *p-value* |
| Gender (female), *n* (%) | 92<br>(72) | 39<br>(46) | 24<br>(56) | 12<br>(33) | 9<br>(47) | 5<br>(23) | 0.0006<sup>a</sup> |
| Age (years), *median*<br>[IQR] | 47<br>[38 - 54] | 58<br>[48 - 67] | 43<br>[33 - 53] | 55<br>[45 - 63] | 46<br>[23 - 52] | 56<br>[49 - 62] | <0.0001<sup>b</sup> |
| Severity, *n* (%) |  |  |  |  |  |  |  |
| Asymptomatic | 12 (9) | --- | 7 (16) | --- | 1 (5) | --- |  |
| Mild | 116 (91) | --- | 36 (84) | --- | 18 (95) | --- |  |
| Hospitalized<br>non-severe | --- | 31 (37) | --- | 9 (25) | --- | 2 (9) |  |
| Hospitalized<br>severe | --- | 41 (49) | --- | 23 (64) | --- | 20 (91) |  |
| Hospitalized<br>(intensive care unit) | --- | 12 (14) | --- | 4 (11) | --- | 0 (0) |  |

IQR: interquartile range (25<sup>th</sup> and 75<sup>th</sup> percentiles),
<sup>a</sup> Chi square test, <sup>b</sup> Mann-Whiney test

### Longitudinal analysis of neutralization activity

All patients were assayed for their plasma neutralization capacity of the original WH1 sequence in a validated pseudovirus assay.^13^ Maximal follow up periods for unvaccinated individuals infected during the first, second and third waves, were 458, 320 and 145 days respectively. In line with previous analyses,^15,17^ irrespective of the infecting virus, hospitalized patients showed a rapid development of neutralizing activity over the first month after symptom onset and a transient decrease reaching a plateau (Figure 1B, 1D, 1F): This was observed only in the first and second wave participants, due to the limited follow up of recently infected patients. In contrast, mildly affected or asymptomatic individuals developed a lower maximal neutralization titer with a flatter behavior (Figure 1A, 1C, 1E), although an early peak could be observed in some of the second wave participants, which had earlier sampling (Figure 1C). Longitudinal analysis using smoothing-splines mixed-effects models showed overlapping kinetics for the different waves in each clinical group (Figure 1G and 1H), although neutralizing activity tended to reach higher values at the peak (around 30 days) in hospitalized patients from the third wave (mostly infected by the alpha variant) (Figure 1H). According to recent data,^23^ we assumed the generation of early short-lived plasmablast/plasma cells and long-lived plasma and memory B cells and modelled data from all patients to a two-phase exponential decay. The longitudinal modeling revealed that hospitalized individuals had a significant rapid first-phase decay (half-life 26 days) and a flat slope in the second phase (half-life 533 days, Figure 1I). Conversely, a flat slope (i.e., not significantly different from 0 in any phase) was observed in individuals with asymptomatic infection or mild disease (Figure 1I). These data confirm that, despite different longitudinal patterns, all infected individuals, outpatients and hospitalized, generate long-lasting neutralizing antibodies.

**Figure 1.**
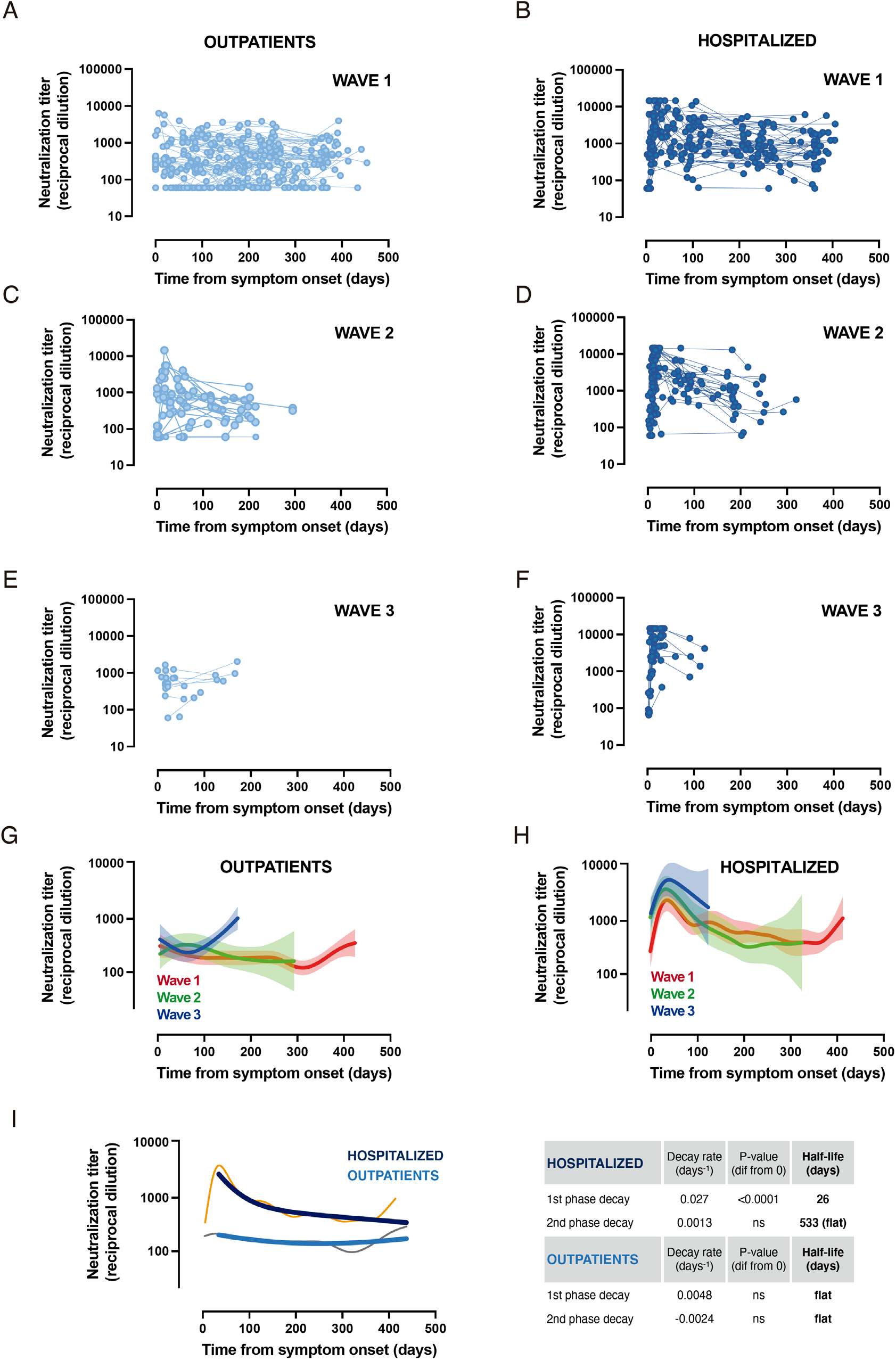
Longitudinal analysis of neutralizing activity. **a-f**, Neutralization titer of 332 individuals according to disease severity (outpatients or hospitalized groups) and date of infection (wave 1 March-June 2020, wave 2 July-December 2020 and wave 3 January-March 2021). Dots are single determinations and lines indicate individual follow up. **g-h**, Longitudinal smoothing-splines mixed-effects models for the different groups are shown in panels a-f. Solid lines indicate the best fit and light areas indicate confidence intervals. **i**, Non-linear models of the full dataset (n=190 for outpatients and n=142 for hospitalized groups) were analyzed by smoothing-splines mixed-effects models (grey and orange narrow lines) or fitted to a non-linear two-phase exponential decay model (light and dark blue lines). Decay rate constants are described on the right side of the figure.

### Impact of vaccination

Massive vaccination campaigns across developed countries have positively impacted the course of the pandemic and have interfered with the follow-up of immune responses induced by natural infection. During routine follow-up visits, we identified 58 vaccinated individuals in our cohort. Participants showed a wide range of vaccination status in terms of type of vaccine (21% received BNT162b2 [Pfizer-BioNTech]), 62% mRNA-1273 [Moderna]) and 17% AZD1222 [AstraZeneca-Oxford]), number of doses (only 55% had received the full 2-dose schedule) and time from the last dose (being BNT162b2 vaccinees analyzed at longer timepoints after vaccination). Despite these differences, vaccines boosted preexisting neutralizing responses in all outpatient (n=40) and hospitalized (n=18) participants (Figure 2A and 2B). A direct comparison of pre- and post-vaccination titers of neutralizing antibodies clearly confirms a highly significant increase in both groups (*p*<0.0001). Pre-vaccination titers tended to be lower in outpatients than hospitalized individuals (*p*=0.0667) and reached comparable levels after vaccination (Figure 2C). However, the heterogeneous vaccine schedules and sampling times prevented further analysis.

**Figure 2.**
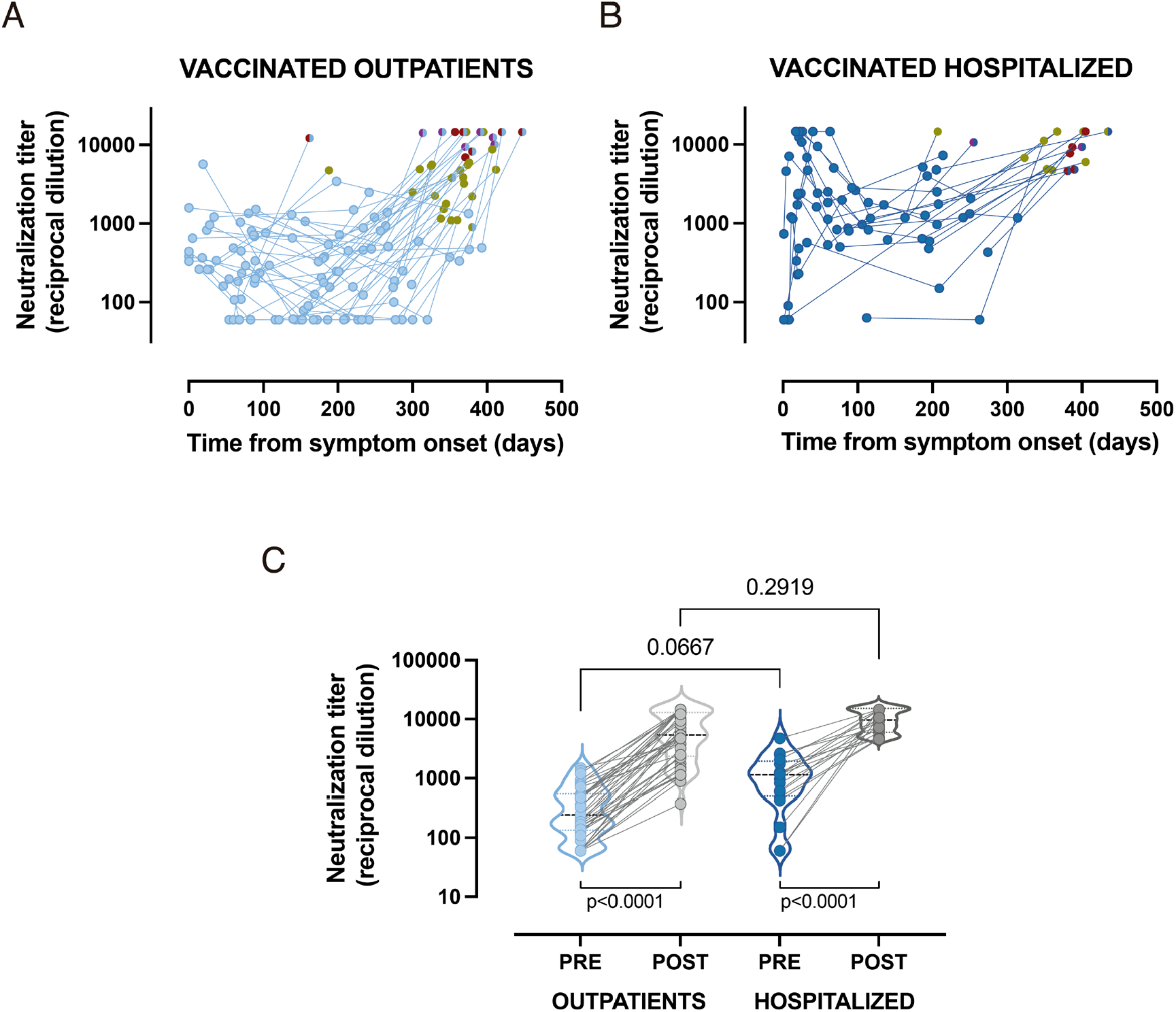
Impact of vaccination on convalescent plasma neutralizing activity. **a-b**, Single measurements (dots) and individual evolution (lines) of the longitudinal analysis for vaccinated mild or asymptomatic (n=40) and hospitalized (n=18) individuals. Blue dots (light or dark, respectively) correspond to pre-vaccination measurements. Post-vaccine data are color coded according to vaccine schedules: BNT162b2 (maroon), mRNA-1273 (red) and AZD1222 (purple); full symbols indicate full schedule (two doses), while half circles indicate one single dose. **c**, Comparison of pre-vaccination and post-vaccination neutralizing antibody titers in both groups. Lower p-values indicate paired comparison (Wilcoxon test) of pre and post values, upper p-value indicates Mann-Whitney comparison of pre-vaccination between groups.

### Impact of viral variants

It is well known that SARS-CoV-2 VOCs show variable degree of resistance to neutralizing responses elicited by natural infection or vaccination. Therefore, to evaluate the impact of two of the most relevant VOCs on long-term neutralizing activity, a subset of 60 unvaccinated individuals with follow-up period beyond 300 days was analyzed. A global analysis showed that long-term neutralizing responses cross-neutralized the WH1 and the alpha (B.1.1.7) variants with similar potency but showed lower titers against the beta (B.1.351) variant (*p*=0.0007, Figure 3A). The subanalysis of groups showed similar but not identical results, we observed a highly significant loss of neutralizing capacity against the beta variant in hospitalized individuals (Figure 3A) but a lower loss in outpatients, reaching significance when compared to the alpha but not to the original WH1 variant (*p*=0.1918, Figure 3A). To quantitatively assess these differences, we compared the ratio of neutralization titer between the tested variants and the original WH1 sequence. These ratios measure the loss of neutralization for each individual and showed no differences between outpatient and hospitalized group for B.1.1.7 variant (with a mean fold change around 1). On the other hand, statistically significant differences were observed for the B.1.351 variant, which induced a higher relative loss of neutralization in hospitalized patients (*p*=0.0350, Figure 3B). As a consequence, even though the median magnitude of neutralization against beta variant was still superior in hospitalized individuals, statistical significance compared to outpatients was lost (Figure 3A).

**Figure 3.**
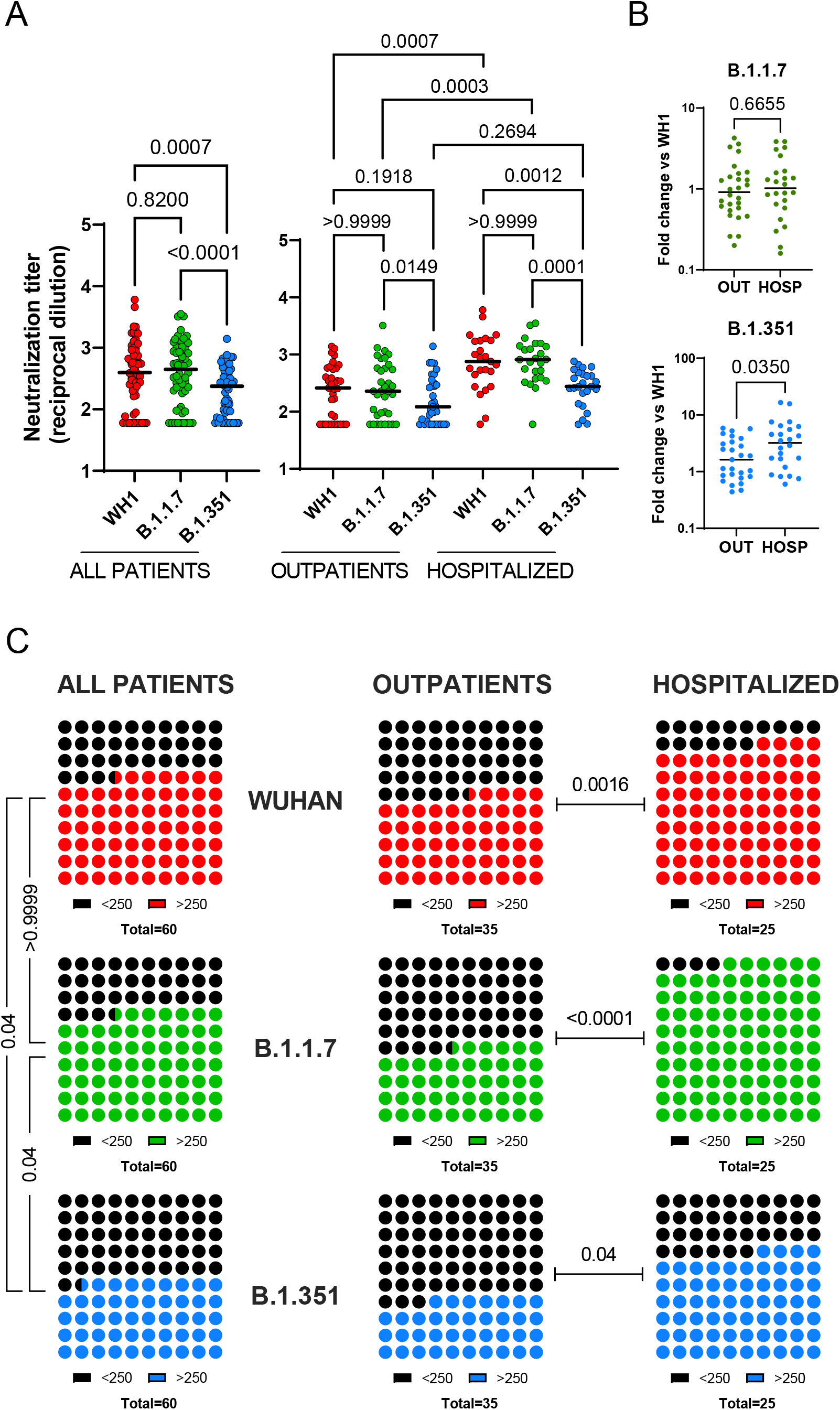
Impact of SARS-CoV-2 variants on long-term neutralizing activity. **a-c**, Neutralization titers, against WH1, B.1.1.7 and B.1.351 spike variants, measured on convalescent plasmas collected more than 300 days after symptom onset from non-hospitalized (n=35) and hospitalized (n=25) patients (Supplementary Table 1). **a**, Neutralization titers (ID50 expressed as reciprocal dilutions) from all the patients (left) or divided into outpatients and hospitalized patients (right). Bars indicate median titer in each group and p-values show the comparison of median titers among the three viruses (Friedman test with Dunn’s multiple comparison) or the comparison of the same variant between the 2 groups (Kruskal-Wallis test with Dunn’s multiple comparison). **b**, Ratios between WH1 and other variants (indicated on top) neutralization titers (lower is better). Bars indicate the median ratio and p-values indicate the comparison of the 2 patient groups (Mann–Whitney test). **c**, Frequency of long-term neutralizers (i.e., individuals with mean neutralizing activity >250 after 300 days post symptom onset) in all patients and separately in hospitalized and non-hospitalized patients. P-values show the comparison of frequency between each variant within all patients and between outpatient and hospitalized for each variant (Chi square test).

Following previous reports correlating protection with neutralization titers,^7^ we estimated the frequency of individuals with high or low neutralization titers using a cutoff value of 250. The analysis showed that 33% of individuals had low neutralization against the WH1 or the alpha variant, increasing to 52% for the beta variant (Figure 3C, *p*=0.0422). In all cases the frequency of low neutralizers was higher in outpatients, reaching a 63% against the beta variant, compared to 36% in hospitalized patients (Figure 3C, *p*=0.0401)

### Factors determining long term neutralizing activity

Despite similar long-term stability in both outpatients and hospitalized individuals, neutralizing activity was highly heterogeneous among individuals, being non-neutralizing and highly neutralizing patients present in both groups (see Figure 3). Therefore, we analyzed the factors that potentially define the magnitude of long-term neutralization in our cohort. A multivariate analysis including severity group, age, gender and infection wave showed that only severity, defined by hospitalization, was independently associated with the magnitude of responses (*p*=0.0285, Figure 4A), while wave (i.e., infecting virus) or gender had no impact (Figure 4B and 4D). Consistent with the close relationship between age and severity, age showed a significant effect in the univariate analysis that was lost in the multivariate model (*p*=0.0951, Figure 4C).

**Figure 4.**
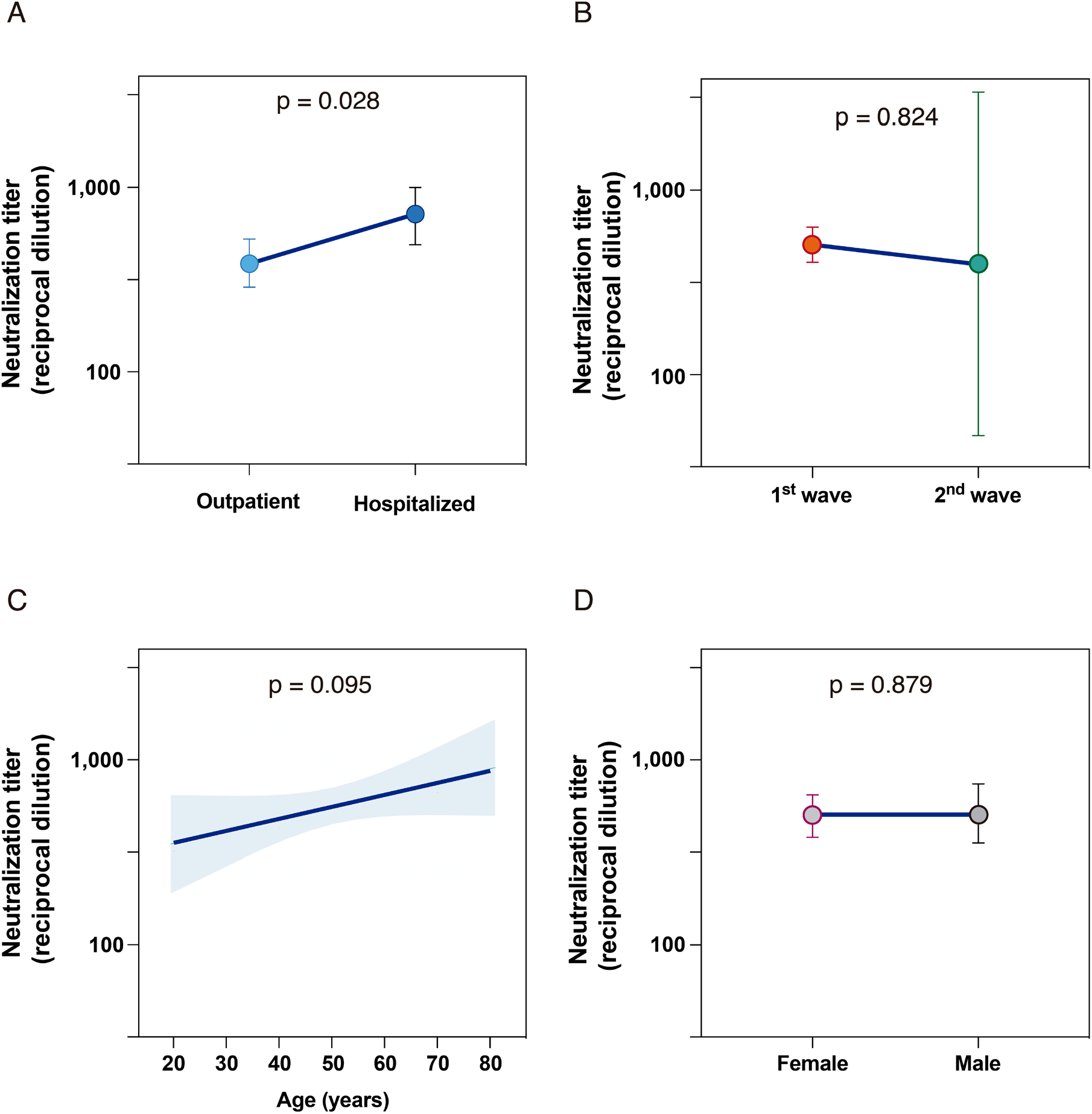
Factors determining long-term neutralizing titer. **a-d**, Factor effects by multivariate linear regression for samples collected more than 300 days post-symptom onset from 99 participants. Estimated effect (dots) and 95% confidence interval (bars or bands) are plotted and the p-value is shown for each predictor covariate: **a**, Severity; **b**, Wave of infection; **c**, Age; **d**, Gender. Multivariate analyses were performed with R-3.6.3 software.

A similar approach was used to assess the impact on beta variant cross-neutralization activity (shown in Figure 3). In this case, the univariate analysis ruled out any impact of age and gender and pointed to severity as the main determinant of reduced neutralization against the beta variant (*p*=0.0259, data not shown).

## Discussion

To our knowledge, this report has analyzed the neutralizing response against SARS-CoV-2 with the longest follow-up to date, with sampling up to month 15 after symptom onset, in a large cohort with a broad spectrum of clinical disease presentation (from asymptomatic to patients requiring intensive care) over different COVID-19 outbreaks in Catalonia. Longitudinal sampling allowed us to model accurate kinetics of neutralizing activity for the different waves (associated with different viral variants). The temporal patterns for each wave appear to repeat themselves independently of the infecting variant, but with a strong impact of disease severity, as previously defined.^13^

In comparison to the apparent short-lasting immunity against seasonal human coronaviruses,^24,25^ the neutralizing response developed against SARS-CoV-2 shows a dynamic pattern similar to the ones described against other coronaviruses that cause severe acute respiratory illness, such as SARS-CoV and MERS-CoV. For these viruses, several studies detected neutralizing antibodies in the first days after diagnosis with a rapid increase peaking between 2 weeks and 1-month post-symptom onset. Thereafter, there was a decline and subsequently a “stabilization” that was maintained beyond 1 year after infection in most cases, and was related to disease severity.^26–32^ Our data demonstrates the long-term (15 months) persistence of neutralizing antibodies against SARS-CoV-2 in most COVID-19 individuals and the fitted model predicts longer stability as it has been described for SARS-CoV and MERS-CoV.^33–38^ This raises an optimistic scenario as neutralizing antibody levels are highly predictive of immune protection,^5,7,39,40^ although sporadic cases of reinfection have been reported, even in the presence of neutralizing antibodies.^41,42^

Our results complement previous studies that evaluated mid-term immunity,^2,17,19,43,44^ being in line with current evidence showing a long-lasting neutralizing response for at least 1 year,^45–47^ the presence of RBD-specific memory B cells^18,48^ and long-lived bone marrow plasma cells.^23^ Although several mechanisms have been proposed that may lead to long-term persistence of antibodies,^49^ the presence of long-lived plasma cells has received more support in recent years^50–52^ and a biphasic model considering short- and long-lived plasma cells has been described.^53,54^ On this basis and considering neutralizing capacity of plasma as a surrogate marker of plasma cell lifespan, we fitted our data to a two-phase exponential decay curve, probably reflecting both short- and long-lived plasma cells. Therefore, our data point to an initial and transient generation or expansion of short-lived SARS-CoV-2 specific plasmablast/plasma cells in hospitalized patients. While selection of high affinity B cells into the germinal centers seems to be a hallmark for the generation of long-lived plasma cells,^55^ short-lived cells can be generated following an extrafollicular response,^51^ which does not necessarily imply immunoglobulin evolution through somatic hypermutation neither selection of high-affinity B cells. Interestingly, hospitalized patients showed a more limited cross-reactive response against B.1.351 variant, suggesting that B cell responses in severe disease, despite being higher in magnitude are poorly cross-neutralizing, probably due to a lower immunoglobulin affinity maturation and limited B cells selection. Although this has been pointed out for early responses in different studies,^2,13,56^ our data extend this observation to the long-term responses, providing evidence for a discordant relationship between magnitude and quality of antibodies in hospitalized individuals.

In the cohort studied, we observed that neutralizing activity is significantly boosted after vaccination, although the longevity of this response still needs to be determined. Based on our data on unvaccinated infected individuals, the vaccination of people who have overcome the SARS-CoV-2 infection should lead to a long-lasting protection. But this information must be interpreted carefully since new emerging variants of the virus could escape both natural and vaccine-induced immunity.^57^

To address the impact of VOCs, we tested neutralization titers against alpha and beta variants. Despite showing lower titers, outpatients demonstrated better cross-neutralization against all variants tested. We also observed a quantitative reduction of titers for the beta variant, resulting in a high frequency of individuals with low (<250) neutralizing capacity that was significantly higher in outpatients. When analyzing the clinical and demographic factors that could influence the long-term neutralizing antibody response, we did not observe any differences between women and men, nor between the first and second infection waves. In contrast, age shows a certain tendency (older participants present higher neutralizing activity) whose significance was evident in the univariate analysis but did not reach significance in the multivariate linear regression. This latter result could indicate that age by itself is not a determinant component, but depends on other cofactors, as could be the severity of the disease, which is highlighted as the main determinant of the magnitude of long-term responses. This is in line with the evidence described so far,^44,47^ although it disagrees with another study describing antibody kinetics influenced by gender.^46^ Despite the clear effect of severity, there is still a high individual heterogeneity in the magnitude of neutralization achieved by participants in each group (outpatients or hospitalized individuals) that needs further study to unveil additional determinants.

Our analysis provides one of the largest datasets on neutralizing activity (in number of participants and follow-up time), but is limited by the lack of parallel data on T-cells and other immune-related factors. In addition, the long-term impact of vaccination is still an open question; therefore, beyond the clear boosting effect observed, we cannot draw further conclusions due to heterogeneous vaccine schedules and sampling times. Our longitudinal analysis confirmed the early decay and long-term maintenance of neutralizing activity observed in other cohorts.^10,12^ Moreover, our data identified different dynamics of short- and long-lived responses after infection. In particular, severity of primary infection is associated with the emergence of short-lived antibodies (not observed in outpatients), and the generation of higher titers of less cross-neutralizing long-lived antibodies (beyond one year).

## Supporting information

STROBE_checklist_cohort

## Data Availability

Further information and requests for resources and reagents should be directed to and will be fulfilled by the Lead Contact, Julià Blanco.
This study did not generate any unique datasets or code.

## Acknowledgements

This work was partially funded by Grifols, the *Departament de Salut* of the *Generalitat de Catalunya* (grant SLD016 to JB and grant SLD015 to JC), the Spanish Health Institute *Carlos III* (Grant PI17/01518 and PI20/00093 to JB; PI18/01332 to JC), CERCA Programme/*Generalitat de Catalunya* 2017 SGR 252, and the crowdfunding initiatives #joemcorono, BonPreu/Esclat and Correos. The funders had no role in study design, data collection and analysis, the decision to publish, or the preparation of the manuscript. EP was supported by a doctoral grant from National Agency for Research and Development of Chile (ANID): 72180406.

We are grateful to all participants and the technical staff of IrsiCaixa for sample processing. Francesc López-SeguÍ provided medical writing support during the preparation of the manuscript.

## Author contributions

JB and BC designed and coordinated the study. EP, BT, SM, FT-F, RO, CR, JR, JV-A, JS and NI-U performed and analyzed neutralization assays. EP, BT and VU performed statistical analysis. MN-J, RP, LM, AC, RT, MM, VG, AV and JC selected patients and coordinated data. JB and EP drafted the manuscript and all authors have made substantial contributions to the revision of the subsequent versions. All authors approved the submitted version of the manuscript and agreed both to be personally accountable for the author’s own contributions and to ensure that questions related to the accuracy or integrity of any part of the work.

## STAR Methods

### RESOURCE AVAILABILITY

#### Lead Contact

Further information and requests for resources and reagents should be directed to and will be fulfilled by the Lead Contact, Julià Blanco.

#### Material availability

The plasmids pcDNA3.4 SARS-CoV-2.SctΔ19 are available upon request to the lead contact.

#### Data and Code Availability

This study did not generate any unique datasets or code.

### EXPERIMENTAL MODEL AND SUBJECT DETAILS

#### Study overview and subjects

The study KING was approved by the Hospital Ethics Committee Board from Hospital Universitari Germans Trias i Pujol (HUGTiP, PI-20-122 and PI-20-217) and was further amended to include vaccinated individuals. All participants provided written informed consent before inclusion.

Plasma samples were obtained from individuals of the prospective KING cohort of the HUGTiP (Badalona, Spain). The recruitment period lasted from March 2020 to March 2021, thus covering the consecutive outbreaks of COVID-19 in Catalonia (Supplementary Figure 1). The KING cohort included individuals with a documented positive RT-qPCR result from nasopharyngeal swab and/or a positive serological diagnostic test. Participants were recruited irrespective of age and disease severity ―including asymptomatic status― in various settings, including primary care, hospital, and epidemiological surveillance based on contact tracing. We collected plasma samples at the time of COVID-19 diagnosis and at 3, 6 and 12 months after diagnosis. Additionally, hospitalized individuals were sampled twice a week during acute infection.

#### Cell lines

HEK293T cells overexpressing WT human ACE-2 (Integral Molecular, USA) were used as target in pseudovirus-based neutralization assay. Cells were maintained in T75 flasks with Dulbecco′s Modified Eagle′s Medium (DMEM) supplemented with 10% FBS and 1 µg/ml of Puromycin (Thermo Fisher Scientific, USA).

Expi293F cells (Thermo Fisher Scientific) are a HEK293 cell derivative adapted for suspension culture that were used for SARS-CoV-2 pseudovirus production. Cells were maintained under continuous shaking in Erlenmeyer flasks following manufacturer’s guidelines.

### METHOD DETAILS

#### Pseudovirus generation and neutralization assay

HIV reporter pseudoviruses expressing SARS-CoV-2 S protein and Luciferase were generated. pNL4-3.Luc.R-.E-was obtained from the NIH AIDS Reagent Program.^58^ SARS-CoV-2.SctΔ19 was generated (GeneArt) from the full protein sequence of SARS-CoV-2 spike with a deletion of the last 19 amino acids in C-terminal,^59^ human-codon optimized and inserted into pcDNA3.4-TOPO. A similar procedure was followed to generate expression plasmids for the alpha and beta variants of SARS-CoV-2 S protein^60^ according to consensus data (www.outbreak.info). Expi293F cells were transfected using ExpiFectamine293 Reagent (Thermo Fisher Scientific) with pNL4-3.Luc.R-.E- and SARS-CoV-2.SctΔ19 (WH1, B.1.1.7 or B.1.351), at an 8:1 ratio, respectively. Control pseudoviruses were obtained by replacing the S protein expression plasmid with a VSV-G protein expression plasmid as reported.^61^ Supernatants were harvested 48 hours after transfection, filtered at 0.45 µm, frozen, and titrated on HEK293T cells overexpressing WT human ACE-2 (Integral Molecular, USA). This neutralization assay has been previously validated in a large subset of samples with a replicative viral inhibition assay.^13^

Neutralization assays were performed in duplicate. Briefly, in Nunc 96-well cell culture plates (Thermo Fisher Scientific), 200 TCID_50_ of pseudovirus were preincubated with three-fold serial dilutions (1/60–1/14,580) of heat-inactivated plasma samples for 1 hour at 37°C. Then, 2×10^4^ HEK293T/hACE2 cells treated with DEAE-Dextran (Sigma-Aldrich) were added. Results were read after 48 hours using the EnSight Multimode Plate Reader and BriteLite Plus Luciferase reagent (PerkinElmer, USA). The values were normalized, and the ID_50_ (reciprocal dilution inhibiting 50% of the infection) was calculated by plotting and fitting the log of plasma dilution vs. response to a 4-parameters equation in Prism 9.0.2 (GraphPad Software, USA).

### QUANTIFICATION AND STATISTICAL ANALYSIS

Continuous variables were described using medians and the interquartile range (IQR, defined by the 25^th^ and 75^th^ percentiles), whereas categorical factors were reported as percentages over available data. Quantitative variables were compared using the Mann-Whitney test, and percentages using the chi-squared test. For the longitudinal analysis of neutralizing activity, patients were grouped into two severity groups according to the WHO progression scale^22^ asymptomatic or mild (levels 1-3), and hospitalized (levels 4-10).

Longitudinal kinetics of neutralization activity for hospitalized and mild groups were analyzed by nonlinear models in two ways, parametric and non-parametric models and stratifying by severity in both cases. We fitted a non-parametric model using smoothing-splines mixed-effects model using the “sme” package of R. The final part of this model, showing an increase in neutralization activity, is unreliable due to the small sample size available in that stretch. We also analyzed the observed decrease of neutralization after 30 days by a biexponential decay model [y=P1*exp(-k1*t) + P2*exp(-k2*t)] fitting a nonlinear mixed-effects model and using “nlme” package of R. In this case three samples were excluded due to their influence in the model fitting since were samples after 350 days with and important increase of neutralization with respect the previous determinations and although we cannot rule out their veracity, they had a great impact on the proper fit of the model due to the lack of sample size in the final part of the follow-up.

Differences in neutralization between both groups after 300 days since symptoms were analyzed. We also analyzed the effect of age and gender using a multivariate linear model adjusting by severity to avoid confusion effects, especially for age that are associated with severity. Statistical analyses were performed using R-3.6.3 (R Foundation for Statistical Computing) and Prism 9.0.2 (GraphPad Software)

## KEY RESOURCES TABLE

| REAGENT or RESOURCE | SOURCE | IDENTIFIER |
| --- | --- | --- |
| **Bacterial and Virus Strains** |  |  |
| pNL4-3.Luc.R-.E- | NIH ARP | Cat#3418 |
| SARS-CoV-2.SctΔ19 | This paper | N/A |
| pcDNA3.4-TOPO | GeneArt/Thermo Fisher Scientific | Cat#810330DE |
| pVSV-G | Clontech | Sánchez-Palomino et al.<sup>61</sup> |
| **Chemicals, Peptides, and Recombinant Proteins** |  |  |
| Fetal Bovine Serum | Thermo Fisher Scientific | Cat#10270106 |
| Dulbecco's Modified Eagle Medium | Thermo Fisher Scientific | Cat#41966052 |
| Expi293 Expression Medium | Thermo Fisher Scientific | Cat#A1435102 |
| Opti-MEM I Reduced Serum Medium | Thermo Fisher Scientific | Cat#31985070 |
| ExpiFectamine 293 Transfection Kit | Thermo Fisher Scientific | Cat#A14524 |
| Versene | Thermo Fisher Scientific | Cat#15040033 |
| Puromycin | Thermo Fisher Scientific | Cat#A1113803 |
| DEAE-Dextran | Sigma-Aldrich | Cat#D9885-100G |
| BriteLite Plus Luciferase | PerkinElmer | Cat#6066769 |
| **Experimental Models: Cell Lines** |  |  |
| Expi293F GnTI- cells | Thermo Fisher Scientific | Cat#A39240 |
| HEK293T/hACE2 cells | Integral Molecular | Cat#C-HA101 |
| **Software and Algorithms** |  |  |
| GraphPad Prism v9.0.2 | GraphPad Software | https://www.graphpad.com/scientific-software/prism/ |
| R v3.6.3 | R Foundation for Statistical Computing | https://www.r-project.org/ |
| "nlme" R Package | R Foundation for Statistical Computing | https://cran.r-project.org/web/packages |
| "sme" R Package | R Foundation for Statistical Computing | https://cran.r-project.org/web/packages |
| **Other** |  |  |
| GeneArt Gene Synthesis | Thermo Fisher Scientific | N/A |

---

**Supplementary Figure 1.**
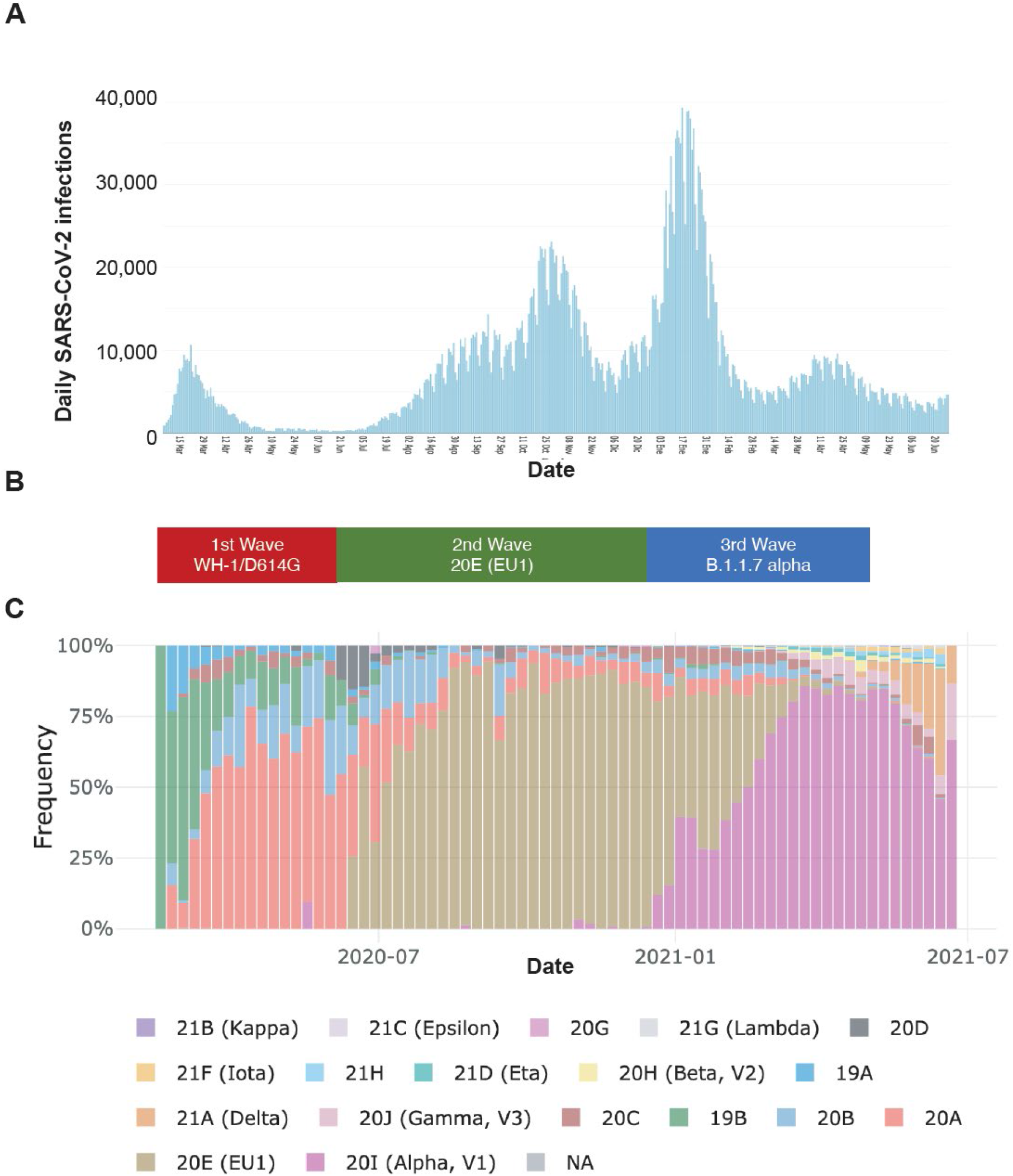
Temporal identification of SARS-CoV-2 waves and main viral variants in Spain. **Panel A** shows the incidence of SARS-CoV-2 infection identifying the main waves between March 2020 and June 2021 (**Panel B**). **Panel C** shows the frequency of circulating SARS-CoV-2 variants in the same time period and clearly associates wave 1 with the 19B and 20A variants, wave 2 with the 20E (EU1) variant and third wave with the 20I (alpha or B.1.1.7) variant.

**SUPPLEMENTARY TABLE 1.** Patients selected for variant analysis (Figure 3)

|  |  | Outpatients | Hospitalized | TOTAL |
| --- | --- | --- | --- | --- |
| Number of cases |  | 35 | 25 | 60 |
| AGE | median [IQR] | 44 [34.5 - 48.5] | 62 [47 - 68] | 46.5 [38 - 57.2] |
| GENDER | Female, n (%) | 28 (80%) | 11 (44%) | 39 (65%) |
| GROUP, n (%) | Asymptomatic | 1 (2.9%) | 0 (0%) | 1 (1.7%) |
|  | Mild | 34 (97.1%) | 0 (0%) | 34 (56.7%) |
|  | Hospitalized non-severe | 0 (0%) | 7 (28%) | 7 (11.7%) |
|  | Hospitalized severe | 0 (0%) | 12 (48%) | 12 (20%) |
|  | Hospitalized (intensive care unit) | 0 (0%) | 6 (24%) | 6 (10%) |

